# Epidemiological characterization of symptomatic and asymptomatic COVID-19 cases and positivity in subsequent RT-PCR tests in the United Arab Emirates

**DOI:** 10.1101/2020.09.23.20200030

**Authors:** Rami H. Al-Rifai, Juan Acuna, Farida Ismail Al Hossany, Bashir Aden, Shamma Abdullah Al Memari, Shereena Khamis Al Mazrouei, Luai A. Ahmed

## Abstract

**Background:** COVID-19 RT-PCR confirmed cases could be symptomatic or asymptomatic. In the United Arab Emirates (UAE), the identified COVID-19 RT-PCR confirmed cases are yet to be characterized. In this study, we characterized the first cohort of COVID-19 RT-PCR confirmed cases reported in the Abu Dhabi Emirate, UAE, according to symptomatic state, and identified factors associated with the symptomatic state. Also, the strength of association between the symptomatic state and testing positive in three subsequent RT-PCR testing rounds was examined and quantified.

**Method:** We analyzed data collected from the first cohort of the RT-PCR confirmed COVID-19 cases reported to the health authorities in the Abu Dhabi Emirate – UAE between February 28 and April 08, 2020. Self-reported sociodemographic, working status, travel history, and chronic comorbidities of 1,249 COVID-19 cases were analyzed according to symptomatic state (symptomatic and asymptomatic). After the first RT-PCR confirmatory test, the results of three subsequent testing rounds were also analyzed.

**Results:** A total of 791 confirmed cases with a mean age of 35.6 years±12.7 (range: 1-81 years) and information on symptomatic state were analyzed. Nearly, 57.0% were symptomatic COVID-19 cases. The most frequent two symptoms were fever (58.0%) and cough (41.0%). The mean age of symptomatic (36.3 year ±12.6SD) was significantly higher than that of asymptomatic cases (34.5 year ±12.7SD). Compared to non-working populations, working in public places (aOR, 1.76, 95% CI: 1.11-2.80), healthcare settings (aOR, 2.09, 95% CI: 1.01-4.31), or in aviation and tourism sector (aOR, 2.24, 95% CI: 1.14-4.40), were independently associated with symptomatic state. Reporting at least one chronic comorbidity was also associated with symptomatic cases (aOR, 1.76, 95% CI: 1.03-3.01). Compared to asymptomatic, symptomatic COVID-19 cases had consistent odds of two or more of testing positive to COVID-19 in three subsequent testing rounds.

**Conclusions:** A substantial proportion of the diagnosed COVID-19 cases in the Abu Dhabi Emirate was asymptomatic. Quarantine of asymptomatic cases along with prevention measures and raising awareness of populations working in high-risk settings is warranted. Further follow up research is needed to understand viral clearance and clinical outcomes according to the symptomatic state of the COVID-19 cases.

## Introduction

Coronavirus disease 2019 (COVID-19) emerged in Wuhan, China, in December 2019.(1) Due to the sustained human-to-human transmission, rapidly, the COVID-19 has spread globally to more than 215 countries affecting over 22 million people with over 777,680 deaths, as of August 16, 2020.(2) About 80% of COVID-19 cases are asymptomatic or mild, 15% are severe requiring oxygen, and 5% are critical infections requiring ventilation.(3) Asymptomatic infection refers to the revealing of nucleic acid of the virus by reverse transcriptase polymerase chain reaction (RT-PCR), in patient not showing typical clinical symptoms.

One of the characteristics that has made COVID-19 pandemic difficult to control has been the infectious status of those who are asymptomatic or very mild symptomatic.(3) Asymptomatic COVID-19 infections have the same infectivity(4) and similar viral load as of symptomatic infections.(5, 6) Contracting viral infection without showing clinical symptoms is highly likely to occur in the event of close contact with confirmed cases. In Boston, 88% of the COVID-19 positive cases were asymptomatic.(7) In Japan, 30.8% of Japanese citizens evacuated from Wuhan were asymptomatic(8) and 51.7% of COVID-19 cases in the “Diamond Princess” were also asymptomatic.(9) The viral RNA can be detected in the respiratory secretions of asymptomatic patients for no less than 3-5 days.(5) Thus, a COVID-19 mild or an asymptomatic case would have the potential of transmitting the virus to other people without any awareness.

The United Arab Emirates (UAE) is described as a melting pot of cultures, with around 85% of the population consisting of expatriates. UAE is burdened with a high prevalence of non-communicable diseases mainly obesity and diabetes mellitus (DM),(10) which may contribute to more susceptibility to the COVID-19. By August 07, 2020, there were over 61,845 COVID-19 cases and 354 deaths reported in the UAE,.(2) Mass active tracing and testing contacts of COVID-19 cases along testing random samples from highly populated residential areas have contributed dramatically in identifying more COVID-19 cases. Relative to the total population, UAE is one of the top 10 countries in terms of the daily number of the tested populations for the COVID-19.(2) In several reported studies, the prevalence of asymptomatic COVID-19 positive cases ranged from 20% to 86% that are defined as individuals with positive viral nucleic acid tests, but without any COVID-19 symptoms.(9, 11-13) To better understand the identified COVID-19 cases, we characterized COVID-19 cases, described factors associated with COVID-19 symptomatic state, and quantified the strength of association between symptomatic state and repeated positivity in subsequent three RT-PCR testing rounds during the disease course.

## Materials and Methods

This study was approved by the Abu Dhabi Health COVID-19 Research Ethics Committee (IRB DOH/CVDC/2020/1518). Owing to the retrospective analysis nature of the study, patient’s informed consent was waived. This study followed and reported according to the Strengthening the Reporting of Observational Studies in Epidemiology (STROBE) reporting guideline.

### Data source

We reviewed the first cohort of 1,249 COVID-19 RT-PCR confirmed cases passively or actively identified and reported to health authorities in the Abu Dhabi emirate up to April 08 2020. The first COVID-19 PCR confirmed case was reported on February 28 2020. Information on the reported symptoms were recorded. Self-reported sociodemographics (age, gender, nationality, place of work), chronic comorbidities (e.g. diabetes mellitus (DM), hypertension, anemia, respiratory diseases), and travel history in the past month, were collected.

#### Specimen collection and viral nucleic acid detection

Upper respiratory tract specimens were collected from nasopharyngeal or oropharyngeal swabs by trained medical staffs. Viral genome detection was performed using RT-PCR according to manufacturer instructions (BGI Genomics Co. Ltd). According to the local health authorities’ guidelines for repeating testing positive cases, RT-PCR testing should be repeated every 48-72 hours. After the first RT-PCR index test, information up to three subsequent RT-PCR testing rounds was available for our COVID-19 cases.

### Evidence synthesis

#### Outcomes of interest

The symptomatic state (presenting symptoms vs not presenting symptoms) of the COVID-19 confirmed cases.

### Exposure variables

We categorized the COVID-19 RT-PCR confirmed cases based on their symptomatic state into asymptomatic or symptomatic (presented with at least one symptom). Age was categorized into four age groups (≤20, 21–39, 40–59, or ≥60 years), to preserve sufficient cases number in each sub-category, nationality regrouped into two sub-groups (Emirati and non-Emirati). Place of work categorized into four sub-groups (not working, working in public places, working in healthcare settings, or working in the aviation and tourism services). This sub-categorization was based on the fact that the risk of exposure to COVID-19 and developing symptoms could be associated to the frequency of being exposed to populations with higher risk of transmitting the viral infection such as travelers from highly infected areas. The “not working” category included housewives, children, visitors, and unemployed people. The “working in public places” category included people working in shopping markets or malls, delivery services, banks, barbershops, hotels, petrol stations, sales, police and security sector, and taxi or bus drivers. Healthcare setting category included healthcare staff (physicians, nurses, ambulance drivers, and para-medics), cleaners, and people working in administrative positions (recipients and cashiers). Aviation and tourism services category included people working in airports, travel agencies, and airport taxi and bus drivers.

The self-reported existence of chronic comorbidities (none or at least one chronic comorbidity) and travel history in the past month (yes or no) each was dichotomized into two subcategories.

### Statistical analysis

Continuous variables are presented as means with standard deviations (SD) and interquartile range (IQR). Categorical variables are presented as frequencies and proportions. The mean difference in continuous variables between symptomatic and asymptomatic COVID-19 cases was compared using Student *t*-test. Difference in proportions of the measured categorical variables between symptomatic and asymptomatic cases was assessed using the Chi-square test.

Crude and multivariable binary logistic regression models were used to quantify the strength of association between the sociodemographic factors and the symptomatic state compared to asymptomatic and the repeated positivity of RT-PCR in the subsequent three testing rounds, among symptomatic compared to asymptomatic cases. Odds ratios (OR) and adjusted OR (aOR) with the 95% confidence intervals were estimated. To control for any potential confounding effect, all measured exposure variables were included in the multivariable model. Collinearity between exposure variables was investigated using the condition index and the variance inflation factor (VIF). There was no collinearity. The maximum VIF value was 1.3 while for the condition index it was 9.6.

The proportions of positive conversion by the symptomatic state were illustrated using Kaplan-Meier plot and the log-rank test was used to determine the difference between the two groups.

Statistical analyses were performed using the IBM SPSS software (version 26). A p-value <0.05 was considered to indicate statistical significance.

## Results

### Profile of the COVID-19 cases

A total of 1,249 RT-PCR confirmed COVID-19 cases were investigated and reported to the health authorities from February 28 to April 08, 2020. There was missing data on the symptomatic state and other key characteristics in 458 cases. These cases were excluded from the analysis. Among the remaining 791 cases, 43.5% were asymptomatic and 56.5% were symptomatic cases (Fig 1).

**Fig 1.**
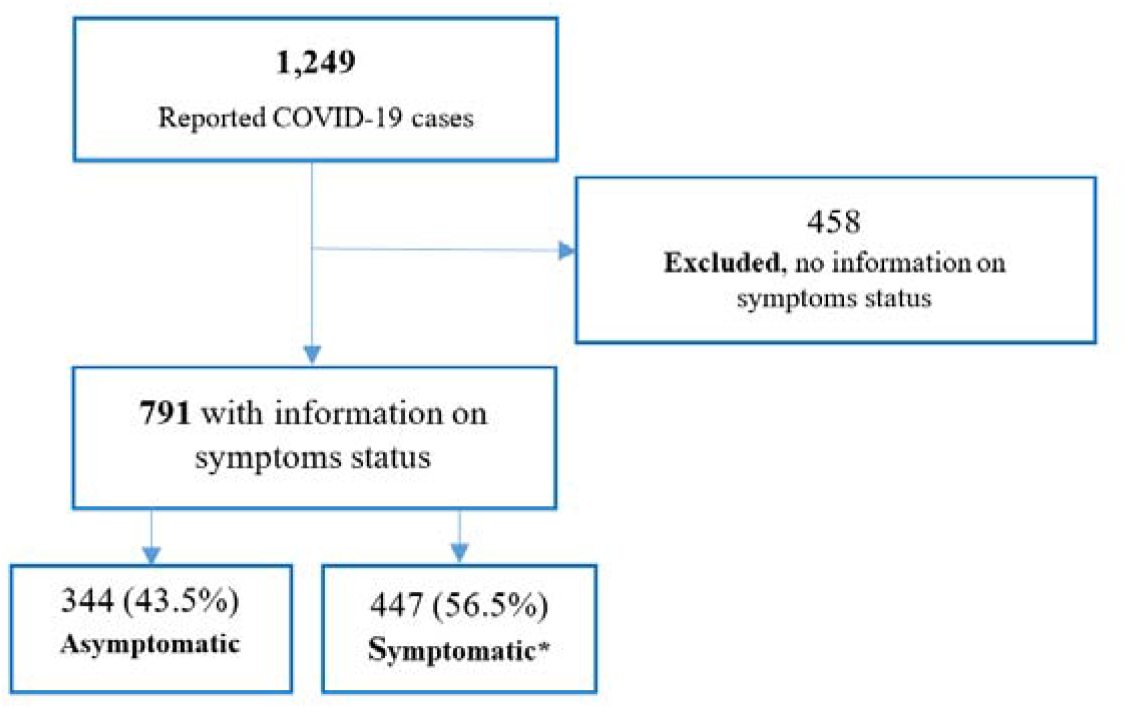
RT-PCR confirmed COVID-19 cases identified from February 28, 2020 to April 08, 2020 in the Abu Dhabi Emirate. * Mean number of symptoms = 1.75 (range 1-6 symptoms)

Of the 447 symptomatic cases, 47.9% presented with only one symptom, 48.5% presented with 2-3 symptoms, and 3.6% presented with four or more symptoms. The most frequent symptoms were fever (58.0%) followed by cough (41.0%), sore throat (18.9%), and headache fatigue (12.4%) (Fig 2).

**Fig 2.**
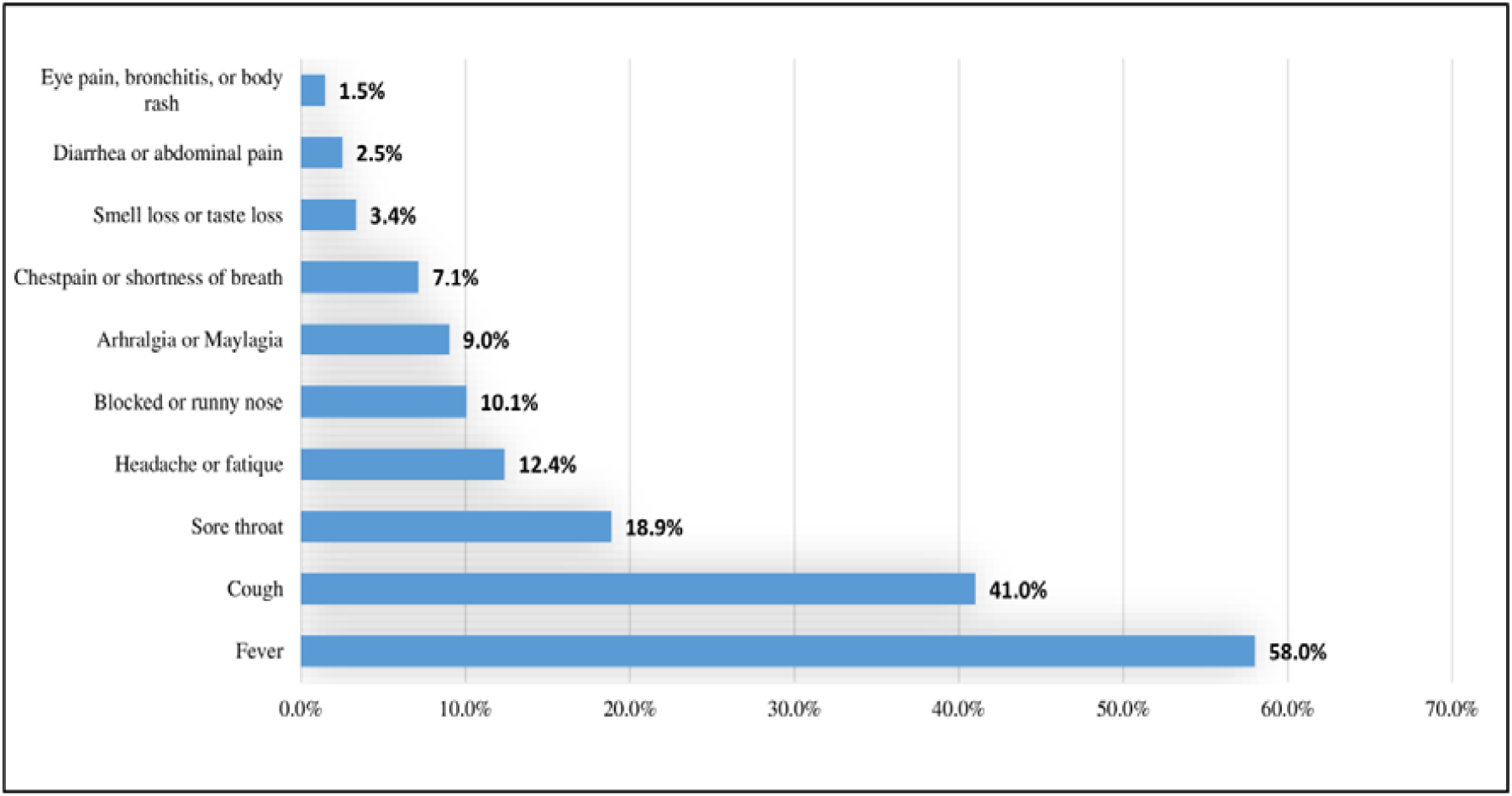
Distribution of symptoms among the 447 symptomatic COVID-19 cases.

The mean age of COVID-19 cases was 35.6 ±12.7 years. Majority of the cases were males (79.2%), 57.6% were Asians, 61.7% were working in public places, 23.6% travelled abroad in the past month, and 13.1% reported having at least one chronic comorbidity, mainly DM (6.0%) and hypertension (5.7%).

Of the 791 COVID-19 cases, 388 (49.1%) were re-tested (second testing round) after an average of 2.7 ±1.4 days from the first testing round. Of these 388 cases, 230 (59.3%) were re-tested (third testing round) after an average of 2.4 ±1.2 days from the second testing round, and of these 230 cases, 70.9% were re-tested (fourth PCR testing) after an average of 3.2 ±1.8 days from the third testing round (Table 1). Overall, the mean number of days from the first to the fourth RT-PCR testing round was 8.3 days ±2.5 days.

**Table 1.**
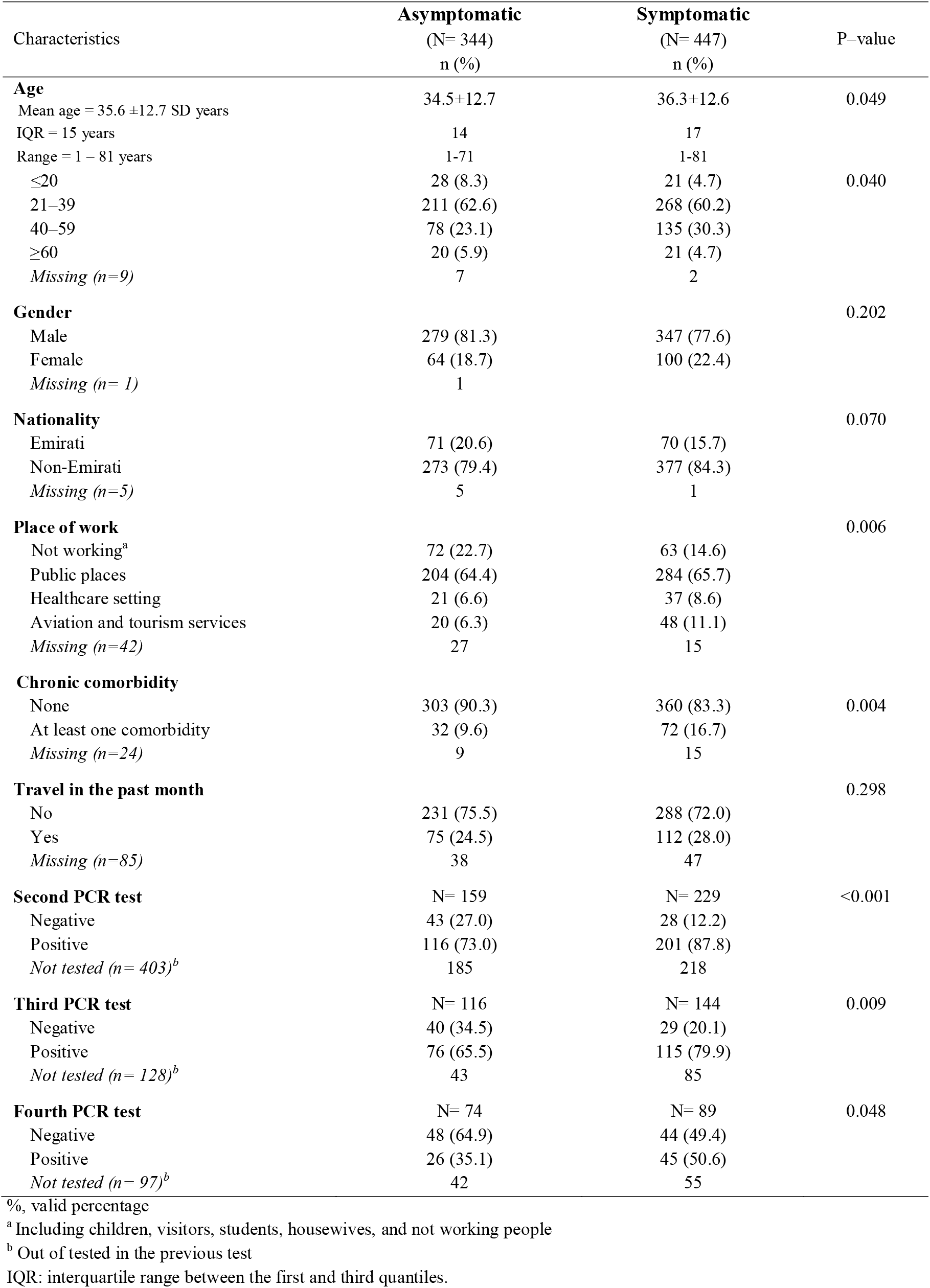
Sociodemographic distribution and subsequent RT-PCR testing outcome of the COVID–19 positive confirmed cases by symptomatic state

| Characteristics | Asymptomatic<br>(N= 344)<br>n (%) | Symptomatic<br>(N= 447)<br>n (%) | P-value |
| --- | --- | --- | --- |
| **Age** |  |  |  |
| Mean age = 35.6 ±12.7 SD years | 34.5±12.7 | 36.3±12.6 | 0.049 |
| IQR = 15 years | 14 | 17 |  |
| Range = 1 – 81 years | 1-71 | 1-81 |  |
| ≤20 | 28 (8.3) | 21 (4.7) | 0.040 |
| 21–39 | 211 (62.6) | 268 (60.2) |  |
| 40–59 | 78 (23.1) | 135 (30.3) |  |
| ≥60 | 20 (5.9) | 21 (4.7) | |
| Missing (n=9) | 7 | 2 |  |
| **Gender** |  |  | 0.202 |
| Male | 279 (81.3) | 347 (77.6) |  |
| Female | 64 (18.7) | 100 (22.4) |  |
| Missing (n= 1) | 1 |  |  |
| **Nationality** |  |  | 0.070 |
| Emirati | 71 (20.6) | 70 (15.7) |  |
| Non-Emirati | 273 (79.4) | 377 (84.3) |  |
| Missing (n=5) | 5 | 1 |  |
| **Place of work** |  |  | 0.006 |
| Not working<sup>a</sup> | 72 (22.7) | 63 (14.6) |  |
| Public places | 204 (64.4) | 284 (65.7) |  |
| Healthcare setting | 21 (6.6) | 37 (8.6) |  |
| Aviation and tourism services | 20 (6.3) | 48 (11.1) |  |
| Missing (n=42) | 27 | 15 |  |
| **Chronic comorbidity** |  |  |  |
| None | 303 (90.3) | 360 (83.3) | 0.004 |
| At least one comorbidity | 32 (9.6) | 72 (16.7) |  |
| Missing (n=24) | 9 | 15 |  |
| **Travel in the past month** |  |  | 0.298 |
| No | 231 (75.5) | 288 (72.0) |  |
| Yes | 75 (24.5) | 112 (28.0) |  |
| Missing (n=85) | 38 | 47 |  |
| **Second PCR test** | N= 159 | N= 229 | <0.001 |
| Negative | 43 (27.0) | 28 (12.2) |  |
| Positive | 116 (73.0) | 201 (87.8) |  |
| Not tested (n= 403)<sup>b</sup> | 185 | 218 |  |
| **Third PCR test** | N= 116 | N= 144 | 0.009 |
| Negative | 40 (34.5) | 29 (20.1) |  |
| Positive | 76 (65.5) | 115 (79.9) |  |
| Not tested (n= 128)<sup>b</sup> | 43 | 85 |  |
| **Fourth PCR test** | N= 74 | N= 89 | 0.048 |
| Negative | 48 (64.9) | 44 (49.4) |  |
| Positive | 26 (35.1) | 45 (50.6) |  |
| Not tested (n= 97)<sup>b</sup> | 42 | 55 |  |

%, valid percentage
<sup>a</sup> Including children, visitors, students, housewives, and not working people
<sup>b</sup> Out of tested in the previous test
IQR: interquartile range between the first and third quantiles.

### Symptomatic versus asymptomatic

The mean age of symptomatic was significantly higher than that of asymptomatic cases (mean difference = 1.8 years, p=0.049). There were more symptomatic (35.1%) aged ≥40 years than asymptomatic cases (29.0%). There were significant differences in the frequency distributions of the place of work between the symptomatic and asymptomatic cases (p=0.006). More than two-thirds of symptomatic and asymptomatic (64.4% and 65.7%, respectively) cases were working in public places. Among the COVID-19 cases working in healthcare settings and aviation or tourism services, there were more symptomatic (19.7%) compared to asymptomatic cases (12.9%). The proportion of symptomatic cases with at least one chronic comorbidity was significantly higher than that of asymptomatic cases (16.7% vs 9.6%, respectively). In each of the three subsequent RT-PCR testing rounds, the proportion of testing positive to COVID-19 was substantially higher among symptomatic compared to asymptomatic cases (Table 1).

### Crude and independent factors associated with symptomatic state

The COVID-19 cases aged 40-59 years had 2.3-fold (95% CI: 1.23-4.34) higher odds to be symptomatic. This association retained insignificant in the multivariable model. Compared to “not working”, working in public places (aOR: 1.76, 95% CI: 1.11–2.80), in healthcare setting (aOR: 2.09, 95% CI: 1.01–4.31), or in aviation and tourism services (aOR: 2.24, 95% CI: 1.14–4.40) was associated with a higher likelihood of having a symptomatic state. Reporting at least one chronic comorbidity was associated with 1.76-time higher likelihood of having symptomatic state (aOR: 1.76, 95% CI: 1.03–3.01). Overall, working compared to not working was associated with 80% increased odds of presenting at least one symptom to COVID-19 (aOR: 1.80, 95% CI, 1.16–2.79) (Table 2).

**Table 2.**
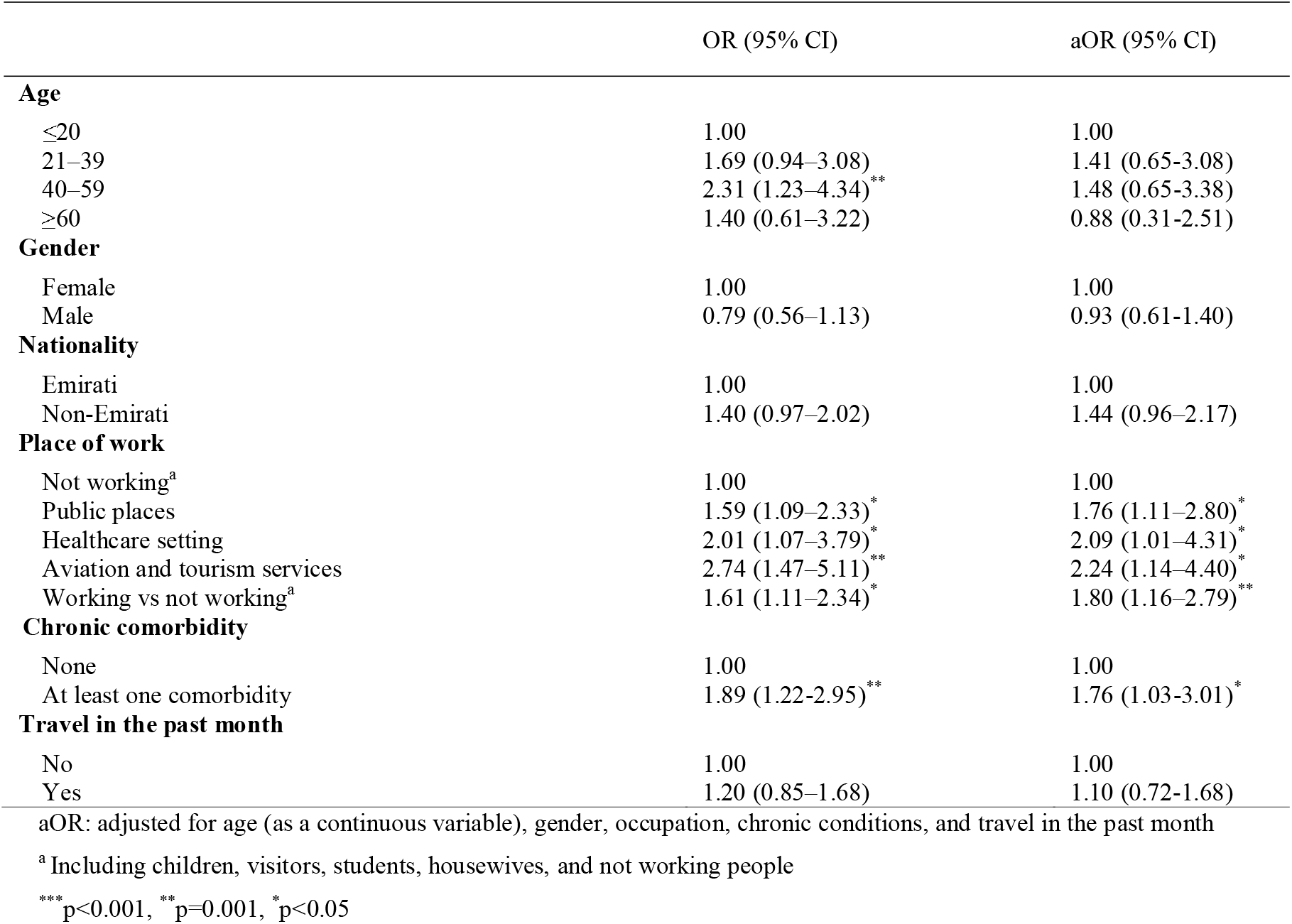
Factors associated with symptomatic state of the COVID–19 RT-PCR confirmed cases compared to asymptomatic cases

### Positivity in subsequent RT-PCR testing rounds

Symptomatic state was associated with prolonged duration of viral shedding compared to asymptomatic state (p-value for log-rank test 0.0026) (Fig 3).

**Fig 3:**
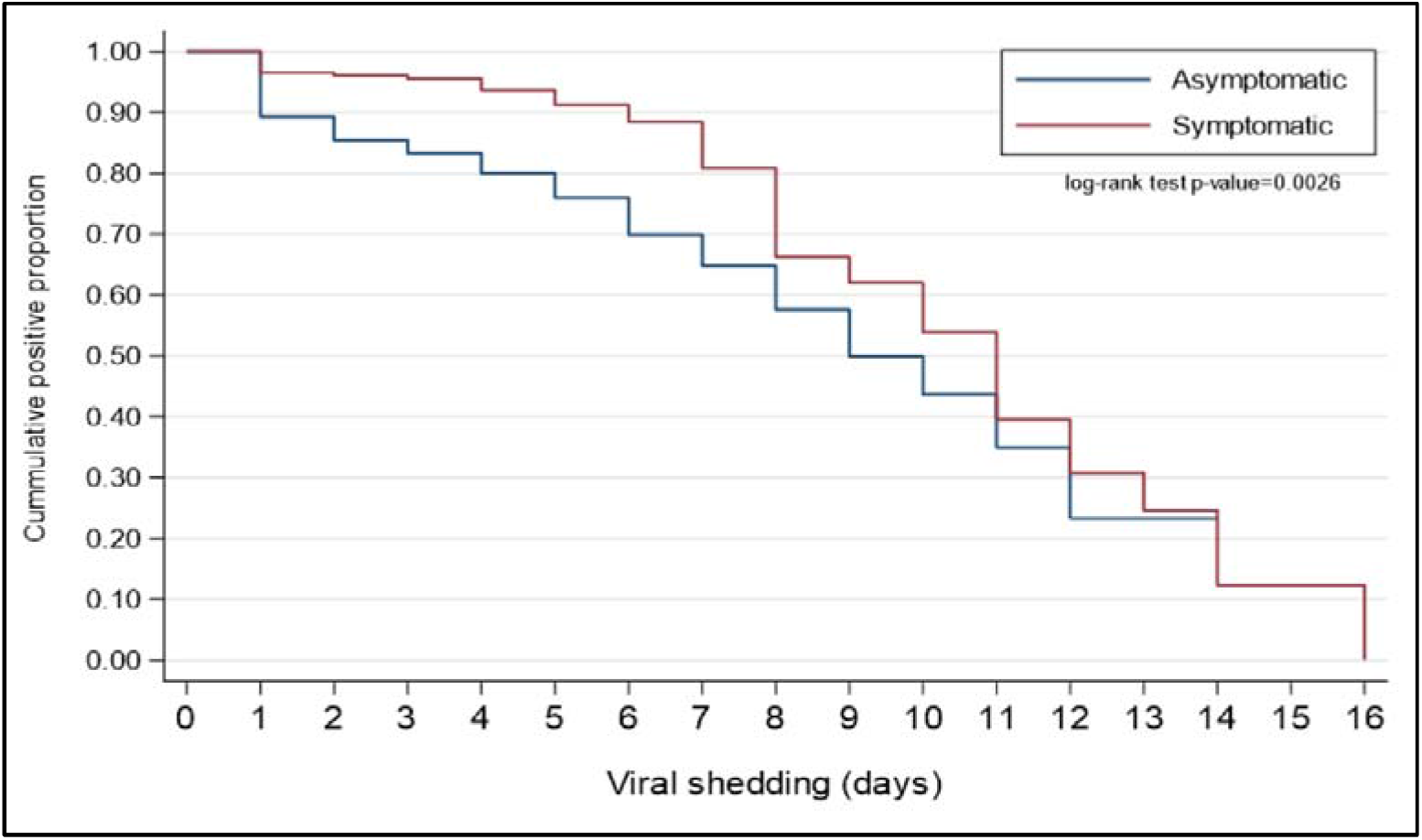
Kaplan-Meier curves for the duration of viral shedding among asymptomatic and symptomatic patients.

In each of the three subsequent RT-PCR testing rounds, symptomatic COVID-19 cases were over two times at higher likelihood of testing positive compared to asymptomatic cases, after adjustment for the potential confounders and difference in the mean day duration between any two subsequent testing rounds (Table 3).

**Table 3.** Crude and adjusted association between symptomatic state and testing positive to COVID–19 in three subsequent RT-PCR testing rounds

| Symptomatic state | Second PCR test |  | Third PCR test |  | Fourth PCR test |  |
| --- | --- | --- | --- | --- | --- | --- |
|  | OR<br>(95% CI) | aOR<br>(95% CI) | OR<br>(95% CI) | aOR<br>(95% CI) | OR<br>(95% CI) | aOR<br>(95% CI) |
| Asymptomatic | 1.00 | 1.00 | 1.00 | 1.00 | 1.00 | 1.00 |
| Symptomatic | 2.66<br>(1.57–4.51)<sup>***</sup> | 2.70<br>(1.45–5.03)<sup>**</sup> | 2.09<br>(1.19–3.65)<sup>*</sup> | 2.26<br>(1.17–4.36)<sup>*</sup> | 1.89<br>(1.00–3.56)<sup>*</sup> | 2.34<br>(1.07–5.14)<sup>*</sup> |

OR: odds ratio
aOR: adjusted odds ratio for the difference in duration (in days) between the two subsequent PCR tests (as a continuous variable), age (as a continuous variable), gender, nationality, number of symptoms, chronic conditions, place of work, and travel in the past month.
<sup>\*\*\*</sup> $p < 0.001$ , <sup>\*\*</sup> $p = 0.001$ , <sup>\*</sup> $p < 0.05$

## Discussion

The findings described COVID-19 RT-PCR confirmed cases according to their symptomatic sate and identified sociodemographic characteristics associated with presenting symptoms in COVID-19 patients. The study also quantified the strength of association between testing positive in three subsequent RT-PCR testing rounds, after the first test. Elder ages, working, or having at least one chronic comorbidity were independently associated with developing symptoms to COVID-19. Symptomatic COVID-19 cases were independently more likely to test positive to COVID-19 after an average duration of 8.3 days from the first confirmatory testing round.

The finding of 43.5% of the reported COVID-19 cases were asymptomatic is higher than the previously reported asymptomatic proportion in Columbia (12.0%),(14) Diamond Princess cruise in Japan (18.0%),(9) Republic of Korea (29.4%),(6) and Japanese evacuated from Wuhan (31.0%).(8) The observed higher proportion of asymptomatic COVID-19 cases in the UAE is merely attributed to the expanded active tracing and screening activities implemented by health authorities. As an outbreak containment strategy, a large number of contacts of the COVID-19 confirmed cases were traced and tested. The first reported two COVID-19 symptomatic cases on February 28, 2020 were for two members of an Italian cycling racing team who had arrived to the UAE on February 20, 2020. For these two cases, a total of 693 individuals were traced and tested for COVID-19 within 1-2 days after identification. Contact tracing and testing could detect a substantial number of potential silent transmitters. The mass tracing and testing activities implemented by the health authorities in the UAE might have contributed to the identification of large number of COVID-19 carriers while they are still in the pre-symptomatic phase. Relative to the total population, UAE is one of the top countries with regard to the total number of performed COVID-19 screening tests. By August 6^th^ 2020, over half (53.2%) of the total population in the UAE were tested for COVID-19.(2)

The crude finding of an inverse association between young age and presenting symptoms to COVID-19 is consistent with a previous report.(15) Younger people are more likely to be healthier with no underlying co-morbidities such as diabetes or hypertension. However, the disappearance of this significant finding in the multivariate model is due to the confounding effect of the “working status”. Younger populations are more likely to be engaged in income-producing activities rather than being retired or unemployed. In the present study, COVID-19 symptomatic state was associated with working in public places (e.g. petrol stations and shopping malls), healthcare settings, or aviation and tourism sector (e.g. cabin crews and airport taxi drivers). Exposure and repeated exposure to COVID-19 carriers are more likely to occur in working places with more exposure to the public. Similarly, working in closed spaces including healthcare settings or in aviation and tourism services potentially increases the risk of exposure and re-exposure to the virus carriers such as confirmed patients and international travelers. Re-exposure to more COVID-19 carriers or contaminated physical services may have contributed to an increased level of the contracted viral load, which may have expedited the symptomatic state.

COVID-19 positive cases with at least one chronic comorbidity were also positively associated with symptomatic state. In our COVID-19 population, DM and hypertension were the most frequent two chronic comorbidities. This observation is in line with other reports that documented DM and hypertension as the most distinctive comorbidities in COVID-19 patients.(16-19) People with chronic comorbidities are more likely to be older in age and to suffer from immune system impairments. DM patients suffer from a lack of energy supply to immune cells that subsequently increases the virulence of infectious microorganisms.(20-22) These impairments weaken the immune system response to microorganisms.(23)(24) Hence, individuals with DM are more likely to be infected and are at a higher risk for complications and death from COVID-19 and other pathogens including SARS and MERS,(17, 25-27) and tuberculosis.(22) A recent study has discussed the mechanism by which DM modulates the host-viral interactions and host-immune responses.(28) Following uptake of COVID-19 by DM patients, the virus invades the respiratory epithelium and other target cells through binding to cell surface angiotensin-converting enzyme 2 (ACE2), and due to the impairment of early recruitment and function of neutrophils and macrophages in DM patients, delay in the initiation of adaptive immunity and dysregulation of the cytokine response may lead to the initiation of cytokine storm that is associated with the symptomatic state and death among DM patients. However, in patients with hypertension, antihypertensive drugs such as ACE inhibitors and statins upregulate ACE2. Increased expressions of ACE2 may favor increased cellular binding of COVID-19.(29-31). Hypertension and DM not only associated with progressing to the symptomatic state, but also delay the viral clearance in COVID-19 patients.(32)

In our study, although the symptomatic state was associated with a delay in testing negative in the subsequent testing rounds compared to asymptomatic cases, a substantial proportion of asymptomatic cases were also tested positive. In the second RT-PCR testing round, after an average of 2.7 days post the first test, almost three quarters (73.0%) while over a one-third (35.1%), after an average of 8.3 days post the first test, of the asymptomatic COVID-19 cases tested positive. This finding of paramount public health significance and implication related to early screening, detection, and timing of quarantine of the asymptomatic COVID-19 cases. Asymptomatic and symptomatic COVID-19 patients were reported to have a similar viral load after a median follow up time of 24 days from diagnosis.(6) Early screening and detection of silent transmitters in the community along evidence-based quarantine timing would positively contribute to controlling the role of silent transmitters on the current and future pandemic waves.

### Limitations and Strengths

Potentially, there are several limitations that should be practiced when interpreting our findings. First, the collected information on the measured sociodemographic and medical conditions was based on self-reports. This potentially may have introduced a risk of reporting bias that could have underestimated or overestimated our findings. However, this is less likely to occur as all the data was collected by trained healthcare staff who are familiar with data collection and with the definition of chronic comorbidities. Second, categorization of the COVID-19 cases into the symptomatic and asymptomatic state without further categorization into a mild, moderate, severe, or critical state. This was because of the lack of other clinical parameters that are necessary for such categorization. However, performing further analysis according to the number of symptoms (asymptomatic, one symptom, 2-3 symptoms, and ≥4 symptoms) was consistent with current findings apart from the significant reduction in the power of the obtained estimates (data not shown). Another important limitation that would potentially further limit the generalizability of our findings is the substantial proportion (36.7%) of missing data on the symptomatic state of COVID-19 cases. Nevertheless, to exclude the potential effect of the missing data, the COVID-19 cases included in this analysis were similar to those with missing data, according to the measured exposure variables except for the number of chronic comorbidities. This difference in the proportion of COVID-19 cases with at least one medical condition could be explained by the observed large proportion of missing data on the self-reported medical conditions of the excluded cases. Also, there was no information on the number of pre-symptomatic patients who subsequently developed symptoms during observation. Finally, all the included studies relied on RT-PCR testing without further investigation using chest computed tomography (CT) imaging. The effect of this could be bi-directional, either overestimation or underestimation, as some cases might have been missed due to false negative result, or some cases might have been included due to the false positive testing to COVID-19.(33, 34)

Despite these limitations, to the best of our knowledge, this is the first study to characterize COVID-19 positive cases in the UAE according to the symptomatic state and to shed light on the strength of association between sociodemographic characteristics and chronic comorbidities with symptomatic state of the COVID-19 cases. Furthermore, our study was based on data collected by trained personal using a standardized data collection and RT-PCR testing procedure. Our study also provided empirical evidence on the positive association between symptomatic state and delay in testing negative compared to asymptomatic COVID-19 cases.

## Conclusions

A substantial proportion of the COVID-19 cases in the Abu Dhabi emirate identified between February 28 and April 08, 2020 was asymptomatic. Working in settings with a higher likelihood of exposure and re-exposure to confirmed or potential virus carriers, or suffering from at least one chronic comorbidity, were independently positively associated with the symptomatic state of the COVID-19 cases. The estimated proportion of asymptomatic cases is a vital parameter for future studies while the estimated strength of association between identified exposures and the symptomatic state and between testing positive in further RT-PCR testing rounds is vital for public health prevention and control interventions. Further follow up cohort studies are needed to have more insight about viral clearance and clinical outcomes in COVID-19 symptomatic and asymptomatic cases.

## Supporting information

STROBE Checklist

## Data Availability

Upon justifiable requests along with a permission from the IRB committee, dataset is available from the corresponding author Dr. Rami H. Al-Rifai.

## Acknowledgment

The authors greatly appreciate efforts of all front liners who contributed to the data and specimen collection.

## Funding

This work received no specific fund.

## Competing interests

None to disclose.

## Author Contributions

All authors contributed to study design and conceptualized the research objectives. RHA, JA, and LAA performed data analysis and interpretation. RHA, drafted the manuscript. All authors reviewed and agreed on the final manuscript.

## Notes

### Competing Interest Statement

The authors have declared no competing interest.

### Clinical Trial

NA

### Author Declarations

This study was approved by the Abu Dhabi Health COVID-19 Research Ethics Committee (IRB DOH/CVDC/2020/1518).

### Summary of Updates

This version revised to correct the name of the last author from "Luai A. Ahemd" to " Luai A. Ahmed".

